# Oral levosulpiride adjuvant to intravitreal ranibizumab for diabetic macular oedema: A 24-week randomized placebo-controlled trial

**DOI:** 10.64898/2026.07.04.26357144

**Authors:** Elva Adán-Castro, Carlos D. Núñez-Amaro, Julian Villarreal, Ilse H. Islas, Andrea Hernández-Quijano, Braulio E. Rodríguez-Chagoya, Marlon García-Roa, Ellery López-Star, Renata García-Franco, Ma. Ludivina Robles-Osorio, Gonzalo Martínez de la Escalera, Carmen Clapp

## Abstract

**Background/Objective:** Diabetic macular oedema (DMO) is a leading cause for visual impairment primarily managed with intravitreal anti-VEGF agents such as ranibizumab (RBZ). Levosulpiride (LSP), a prokinetic medication, was recently repositioned as a safe oral treatment for naive DMO. Here, we investigated the adjuvant effect of oral LSP in combination with intravitreal RBZ injections for treating persistent DMO.

**Subjects/Methods:** Double-blinded, dual-centre, phase 2 trial in patients with centre-involving DMO randomly assigned to be orally treated with placebo (15 patients, 18 eyes) or LSP (18 patients, 19 eyes) along with 3 successive (4 weeks apart) RBZ intravitreal injections and a 24-week follow-up.

**Results:** Baseline best-corrected visual acuity (BCVA) improved (p≤0.04) at week 12 in both RBZ+placebo and RBZ+LSP, but improvement was maintained (p=0.009) at week 24 only in RBZ+LSP. In agreement, longitudinal changes from baseline in BCVA from weeks 12 to 24 defined superior (p=0.02) visual gains measured by the Area Under the Curve (AUC) in RBZ+LSP vs. RBZ+placebo. The baseline value of mean central foveal thickness (CFT) decreased (p≤0.002) in both groups at week 12 and CFT reduction was significant (p=0.006) at week 24 only in RBZ+LSP. Also, longitudinal changes from baseline in CFT resulted in a higher AUC reduction (p≤0.04) at weeks 4 to12 in RBZ+LSP vs. RBZ+placebo. No significant adverse side effects were detected.

**Conclusions:** Adjunctive LSP showed functional and anatomical benefits over the first-line therapy with RBZ. Adjuvant properties may involve the LSP-induced intraocular upregulation and downregulation of vasoinhibin and VEGF, respectively. Larger clinical trials are warranted.

## INTRODUCTION

Diabetic macular oedema (DMO) is a major cause of visual impairment worldwide. It is characterized by leakage from retinal capillaries and an overproduction of vascular endothelial growth factor (VEGF), which contributes to retinal swelling. The primary treatment has become the intravitreal injection of anti-VEGF agents, especially ranibizumab (RBZ), a humanized monoclonal antibody fragment that targets VEGF [1]. However, despite its effectiveness, RBZ treatment has limitations including variable patient response, the need for repeated injections overtime, and potential safety concerns. These challenges have encouraged the development and investigation of adjunctive or alternative therapies to improve treatment outcomes [1–3].

Levosulpiride (LSP), a prokinetic dopamine D2-receptor blocker, was recently repositioned as an effective and safe oral treatment for DMO [4]. Oral LSP for 8 weeks improved visual and retinal structural outcomes in patients with centre-involving DMO with efficacy like intravitreal RBZ [5] by mechanisms including (a) the blockage of dopamine D-2 receptors at the pituitary level causing hyperprolactinemia-induced increase in intraocular vasoinhibin [6,7] a prolactin fragment that inhibits retinal vascular leakage induced by VEGF and diabetes [8,9]; and (b), the downregulation of VEGF and placental growth factor (PlGF) in the eye [4].

Given that LSP increases ocular vasoinhibin [7] and decreases the vitreous concentrations of both VEGF and PlGF [4], we hypothesized that adjunctive oral LSP combined with the intravitreal injection of the VEGF inhibitor RBZ would achieve multimodal suppression of retinal oedema. Such complementary mechanisms may confer additive or synergistic therapeutic effects, resulting in improved anatomical and visual outcomes while reducing treatment burden.

Here, we present a randomized, placebo-controlled study evaluating visual and anatomical outcomes in patients with centre-involving DMO treated with intravitreal RBZ combined with oral LSP over a 24-week follow-up period.

## PATIENTS AND METHODS

This is a phase 2, double-blinded, randomized controlled clinical trial conducted from March 2019 to February 2026 at two sites in Querétaro, Mexico [Instituto Mexicano de Oftalmología (IMO) and Instituto de la Retina del Bajío] and sponsored by Universidad Nacional Autónoma de México (UNAM, grant 405PC), Consejo Nacional de Humanidades, Ciencias y Tecnologías (CONAHCYT, grant A1-S-9620B), and Secretaría de Ciencia, Humanidades, Tecnología e Innovación (SECIHTI, grant CBF-2025-I-908). The study adhered to the guidelines of the Declaration of Helsinki, and the protocol and consent forms were approved by the Bioethics Committees of the Instituto de Neurobiología, UNAM and the IMO. Each subject provided written informed consent, and the study was supervised by an independent data and safety monitoring committee. The study was part of the clinical trial registered at www.clinicaltrials.gov in May 2017 under the identifier NCT03161652. The protocol details including settings and locations, patients and public involvement, eligibility criteria, enrolment and randomization, blinding, outcome measures, sample size estimation, data collection, management and sharing, and safety have been reported [10].

Fifty-six mestizo patients with type 2 diabetes (out of 115 screened) were eligible centre-involving DMO participants [aged 40 to 69 years, best-corrected visual acuity (BCVA) between 55 and 5 Early Treatment Diabetic Retinopathy Study (ETDRS) letters at 4 m (20/20 to 20/200 Snellen equivalent), and central foveal thickness (CFT) of > 260 μm]. After signing informed consent, blood samples were withdrawn to evaluate basal prolactin levels and safety parameters [thyroid stimulating hormone (TSH), glycated haemoglobin, glutamic pyruvic transaminase, and creatinine]. A total of twenty-one patients were excluded from the study for the following reasons: four due to hyperprolactinemia (prolactin levels >20 ng/ml), three because of glomerular filtration rates below 31 mL/min, twelve for having proliferative diabetic retinopathy, one due to unavailability of RBZ, and one who failed to attend the appointment. The remaining 35 subjects were randomized by the study coordinator based on a computer-generated list of random numbers in a 1:1 allocation ratio to receive LSP (DISLEP®, Ferrer Therapeutics; 25-mg orally TID) or placebo (P, lactose pill orally TID) for 24 weeks, in combination with 3 successive (4 weeks apart) intravitreal RBZ injections (0.5 mg/injection). RBZ, LSP, and P treatments initiated simultaneously and study visits were scheduled every 4 weeks. Of the 35 subjects enrolled, 33 (17 females and 16 males) completed the first 12 weeks of treatment (15 in the RBZ+P group and 18 in the RBZ+LSP group). Two participants from the RBZ+P group were removed due to a shortage of RBZ in Mexico. Out of these 33 subjects, 21 (10 in RBZ+P and 11 in RBZ+LSP) completed the full 24-week study. Twelve participants dropped out for the following reasons: three RBZ+P subjects did not adhere to study visits, one RBZ+LSP patient suffered a traumatic retinal detachment, and two RBZ+P plus 6 RBZ+LSP required reinjection between weeks 16 and 20.

Patients were evaluated at baseline and at study visits every 4 weeks using medical history, physical examination, BCVA, optical coherence tomography (OCT), fundoscopy, and laboratory analysis of blood samples. TSH was evaluated only once at baseline, while HbA1c and fluorescein angiography were assessed at baseline, weeks 12, and 24. Both, patients and investigators assessing outcomes were blind to the treatment assignment.

### Primary endpoints

BCVA, OCT, and fluorescein angiography were assessed by certified examiners using standardized protocols as reported [10]. The mean values of BCVA and CFT at baseline (week 0), week 12, and week 24, along with longitudinal analysis comparing the same eye before and after treatment at weeks 4, 8, 12, 16, 20, and 24, served as primary endpoints. These measures reflect the treatments impact over time. Assessments at weeks 12 and 24 included the proportion of eyes demonstrating improved changes from baseline defined as a gain of 5 or more ETDRS letters in BCVA and a reduction of 50 µm or more in CFT.

### Secondary endpoint

Prolactin levels in serum confirmed adherence to LSP treatment (also monitored by counting drug tablet return) and were quantified using the IMMULITE 2000 XPi immunoassay system (Siemens, Munich, Germany). The intra-assay and inter-assay coefficients of variation were less than 1%.

### Safety assessment

The safety of the study medications (RBZ+LSP and RBZ+P) was evaluated through medical history review, physical examination, measurement of ocular pressure, and laboratory blood tests including HbA1c, fasting glucose levels, and creatinine. Additionally, adverse effects were monitored by nondirective questioning of patients during each visit or between visits to capture any reported side effects.

### Statistics

GraphPad Prism Software Inc. version 10.6.1 was used. Data normality was assessed using the Shapiro-Wilk test. Statistical differences between two groups were determined by Student’s t-test or Welch’s t-test when distribution was normal and the Mann-Whitney U-test when not. The chi-square test and Fisheŕs exact tests were used to evaluate differences between categorical variables. Analyses of repeated measurements followed the intention-to-treat principle, with all randomized participants contributing all available longitudinal data. Outcomes were analysed using a mixed-effects model for missing values [11] and multiple comparisons were adjusted using Šidák correction. The threshold for significance was set at p<0.05.

## RESULTS

### DMO patients’ demographics, clinical, and ophthalmic characteristics

Table 1 presents the demographics, clinical, and ophthalmic characteristics of the DMO groups before and at the end of the 24-week study. Thirty-three patients with type 2 diabetes and centre-involving DMO were randomized to receive either oral placebo (n=15) or LSP (n=18) in combination with 3 intravitreal injections of RBZ administered 4 weeks apart. Both groups were well balanced at baseline in terms of demographics (age, sex, and diabetes duration), clinical characteristics [HbA1c, fasting serum glucose, kidney function (serum creatinine and glomerular filtration rate), hepatic function (serum glutamic pyruvic transaminase), ocular pressure], serum prolactin levels, and proportion of patients receiving DMO treatment (more than one intravitreal injection and/or laser photocoagulation) two to nine months prior to study entry [60% vs. 77.78%; p=0.44; odds ratio [OR] = 0.42; 95% confidence interval [CI] = 0.11-1.83] (Table 1). Ophthalmic characteristics at baseline were comparable between the eyes from RBZ+P and RBZ+LSP groups in BCVA (33.65 ± 12.33 vs. 30.74 ± 17.18 number of ETDRS letters read at 4 m; p=0.76) and CFT (404 ± 120.4 vs. 446 ± 133.2 µm; p=0.32). Twenty one participants (10 RBZ+P and 11 RBZ+LSP) completed the full 24-week follow-up. Twelve patients were lost between weeks 16 to 20 due to the following reasons: one RBZ+LSP patient suffered a traumatic retinal detachment, three RBZ+P patients missed visits, and eight underwent RBZ re-injection (six RBZ+LSP and two RBZ+P]. Both groups remained similar at the end of the 24 weeks, except for serum prolactin values that significantly increased in patients receiving RBZ+LSP (Table 1).

**Table 1.** Demographic, clinical and ophthalmic characteristics of the diabetic macular oedema groups.

| Characteristic | BEFORE TREATMENT |  |  | AFTER 24-WEEK TREATMENT |  |  |
| --- | --- | --- | --- | --- | --- | --- |
|  | RBZ + P<br>(15 patients, 18 eyes) | RBZ + LSP<br>(18 patients, 19 eyes) | *p | RBZ + P<br>(10 patients, 12 eyes) | RBZ + LSP<br>(11 patients, 11 eyes) | *p |
| Age, years (SD) | 61.8 (5.39) | 60.5 (6.31) | 0.53<sup>a</sup> |  |  |  |
| Sex F n (%) | 6 (37.5%) | 11 (61.1%) | 0.22<sup>b</sup> |  |  |  |
| DM2 years (SD) | 17.5 (8.30) | 15.9 (7.47) | 0.56<sup>a</sup> |  |  |  |
| HbA1c (SD) | 7.30 (1.44) | 8.05 (2.06) | 0.24<sup>a</sup> | 7.75 (1.70) | 7.46 (2.29) | 0.30<sup>c</sup> |
| Glucose mg/dL (SD) | 144 (54.3) | 154 (67.5) | 0.67<sup>a</sup> | 126 (35.3) | 129 (59.6) | 0.93<sup>c</sup> |
| SCr mg/dL (SD) | 1.01 (0.32) | 0.94 (0.33) | 0.34<sup>c</sup> | 1.01 (0.25) | 1.22 (0.47) | 0.47<sup>c</sup> |
| eGFR mL/min (SD) | 79.3 (20.7) | 82.2 (20.6) | 0.69<sup>a</sup> | 79.8 (18.6) | 66.8 (20.5) | 0.14<sup>a</sup> |
| SGPT U/L (SD) | 21.6 (6.81) | 23.6 (6.30) | 0.47<sup>c</sup> |  |  |  |
| SPRL ng/mL (SD) | 7.83 (3.02) | 8.26 (3.00) | 0.68<sup>a</sup> | 7.79 (2.89) | 119 (58.6) | <0.0001<sup>d</sup> |
| Prior DMO treatment<br>n (%) | 9 (60%) | 14 (77.78%) | 0.44<sup>e</sup> |  |  |  |
| IOP mmHg (SD) | 14.4 (3.43) | 13.7 (3.85) | 0.59<sup>c</sup> | 13.5 (4.25) | 14.6 (2.80) | 0.47<sup>a</sup> |
| BCVA number of ETDRS<br>letters at 4m (SD) | 33.65 (12.33) | 30.74 (17.18) | 0.76<sup>c</sup> | 35.42 (13.10) | 37.36 (14.74) | 0.74<sup>a</sup> |
| CFT µm (SD) | 404.1 (120.4) | 446.2 (133.2) | 0.32<sup>c</sup> | 343.3 (106.9) | 319.45 (109.29) | 0.91<sup>c</sup> |

Values represent the mean of the number (n) of patients (lines 1 to 10) or the mean of the number of eyes (lines 11 to 13).
RBZ ranibizumab; P placebo; LSP levosulpiride; SD standard deviation; DM2 diabetes mellitus type 2; HbA1c glycosylated haemoglobin; SCr serum creatinine; eGFR estimated glomerular filtration rate (CKD-EPI equation); SGPT serum glutamic pyruvic transaminase; SPRL serum prolactin; IOP intraocular pressure; BCVA best corrected visual acuity; CFT central foveal thickness.
\*p vs. respective RBZ+P values based on <sup>a</sup>Student's t test, <sup>b</sup>Chi-square test, <sup>c</sup>Mann–Whitney U test or <sup>d</sup>Welch test and <sup>e</sup>Fisher exact test.

The percentage of patients requiring retreatment before the study’s conclusion was higher in the RBZ+LSP group compared to the control group, but this difference was not statistically significant (33.3% [6/18] vs. 13.3% [2/15], p=0.24; OR = 0.30; 95% CI, 0.056 to 1.75). At the 24-weeks point, additional reinjections were administered to two patients in the RBZ+LSP group and four patients in the RBZ+P group, resulting in comparable retreatment total rate between the groups (44% [8/18] vs. 40% [6/15], respectively).

### BCVA and CFT outcomes

At 12 weeks, the mean number of reading letters from baseline increased significantly in both the RBZ+P (p=0.043) and RBZ+LSP (p=0.015) groups (Fig. 1A). However, at 24 weeks, this visual improvement remained significant (p=0.009) only in the RBZ+LSP group (Fig. 1A). Likewise, a significant reduction in mean CFT was observed at 12 weeks in both the RBZ+P group (p=0.002) and the RBZ+LSP group (p<0.001), and at 24 weeks only the RBZ+LSP group maintained this significant improvement (p=0.006) (Fig. 1B).

**Figure 1.**
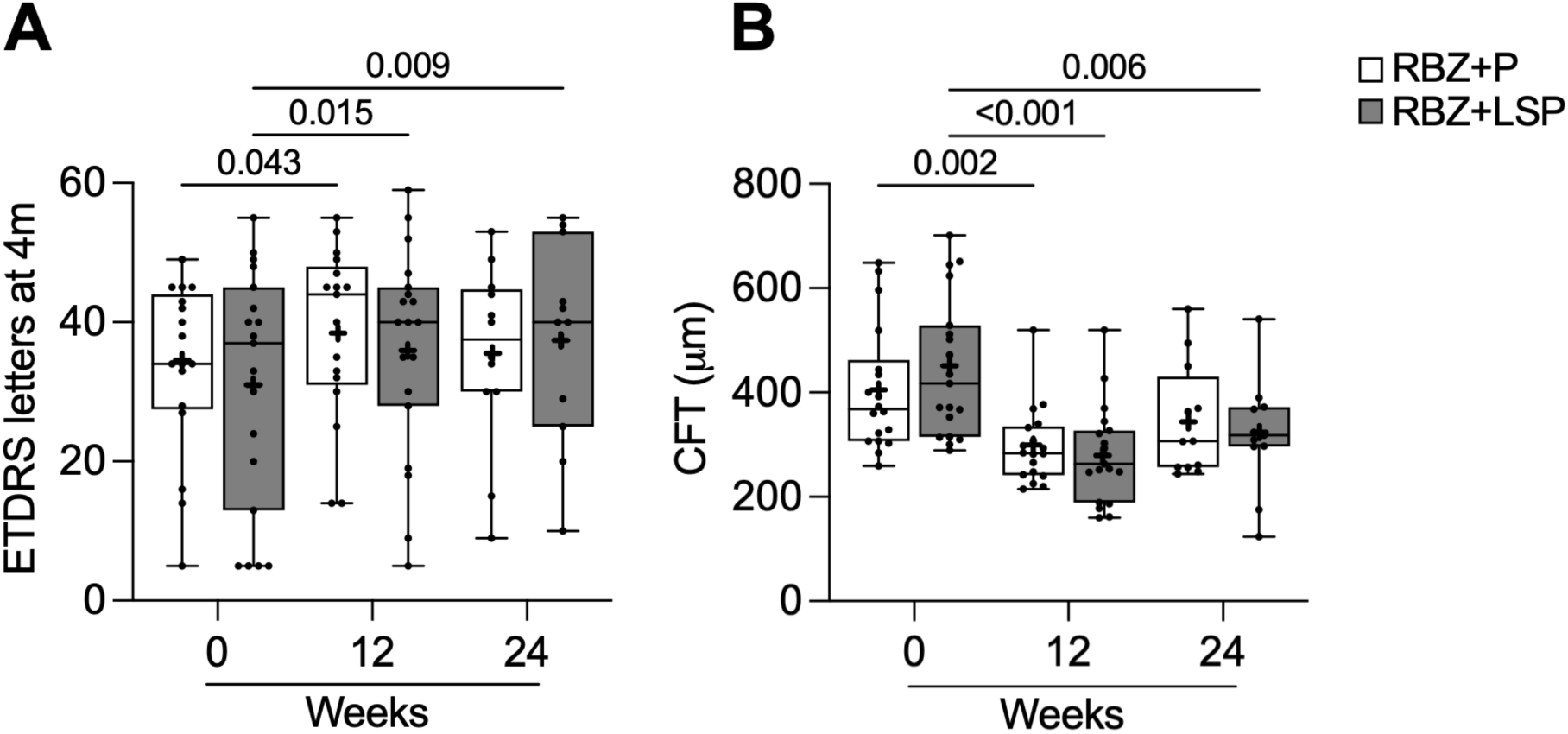
The combination of intravitreal ranibizumab (RBZ) with oral levosulpiride (LSP) but not with oral placebo (P) sustained both anatomical and visual acuity improvements at the 24-week endpoint. Box plots illustrating the distribution of **(A)** the number of ETDRS letters and **(B)** central foveal thickness at baseline (week 0), and at weeks 12 and 24. Data are presented for eyes from patients treated with RBZ combined with either P or LSP. Each box plot displays the median, interquartile range, and minimum and maximum values, with the mean indicated by a plus sign (+) and individual values shown. Sample sizes at weeks 12 and 24 were 18 and 12 eyes, respectively, in the RBZ+P group, and 19 and 11 eyes, respectively, in the RBZ+LSP group. Statistical significance was assessed using a Mixed-Effects Model with Šidák-adjusted multiple comparisons and corresponding p values indicated.

The mean longitudinal increase in BCVA relative to baseline was comparable between groups during the initial 12 weeks of treatment (Figure 2A, B) and, from weeks 16 to 24, the RBZ+LSP group showed superior visual acuity outcomes compared to the RBZ+P group (Fig. 2A, C). Correspondingly, the area under the curve (AUC) for BCVA from baseline to 24 weeks (Figure 2A) and from weeks 12 to 24 (Fig. 2C) revealed a trend toward significance (p=0.07) and a significant difference (p=0.02), respectively, favouring the RBZ+LPS group.

**Figure 2.**
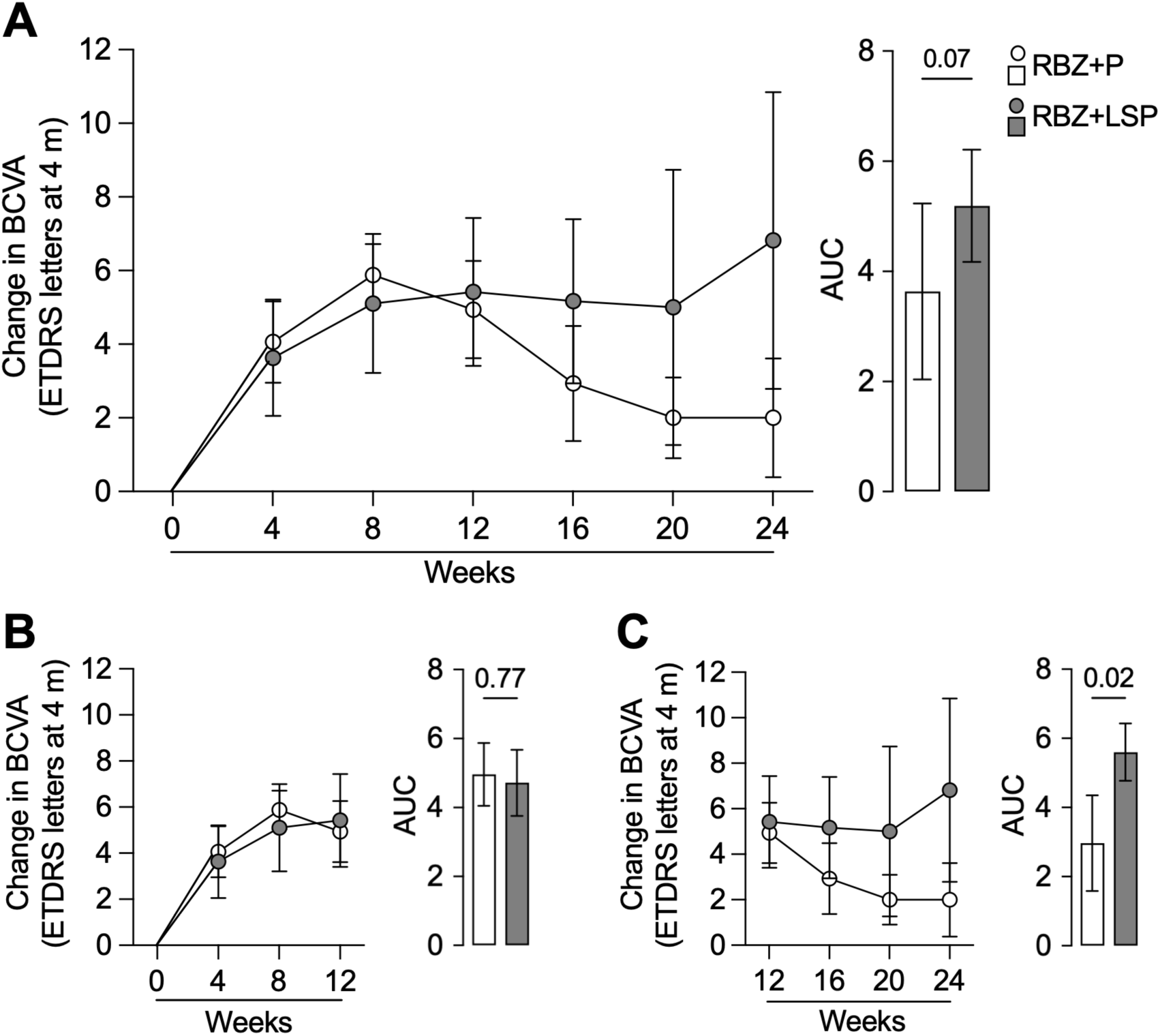
The combination of intravitreal ranibizumab (RBZ) with oral levosulpiride (LSP) improved the change from baseline in best corrected visual acuity (BCVA). Mean longitudinal changes in BCVA are shown for the same eyes at baseline (week 0) and after treatment at weeks 4 to 24 **(A)**, weeks 4 to 12 **(B**), and weeks 12 to 24 **(C)**. Data are presented for eyes from patients treated with RBZ combined with either placebo (P) or LSP. Bars represent the area under the curve (AU) comparing the corresponding groups. Values for the first 12 weeks are expressed as mean ± SEM for 18 and 19 eyes in the RBZ+P and the RBZ+LSP groups, respectively; for the last 12 weeks, values are mean ± SEM for 12 and 11 eyes in the RBZ+P and the RBZ+LSP groups, respectively. AUC values were compared using an unpaired t-test, with p values indicated.

The mean longitudinal reduction of CFT from baseline was consistently greater in the RBZ+LSP group throughout the 24-week period (Fig. 3A), particularly during the first 12 weeks (Fig. 3B). This finding was supported by both a near-significant (p=0.09) and a significant (p=0.02) difference in AUC measurements for the overall and early study periods, respectively.

**Figure 3.**
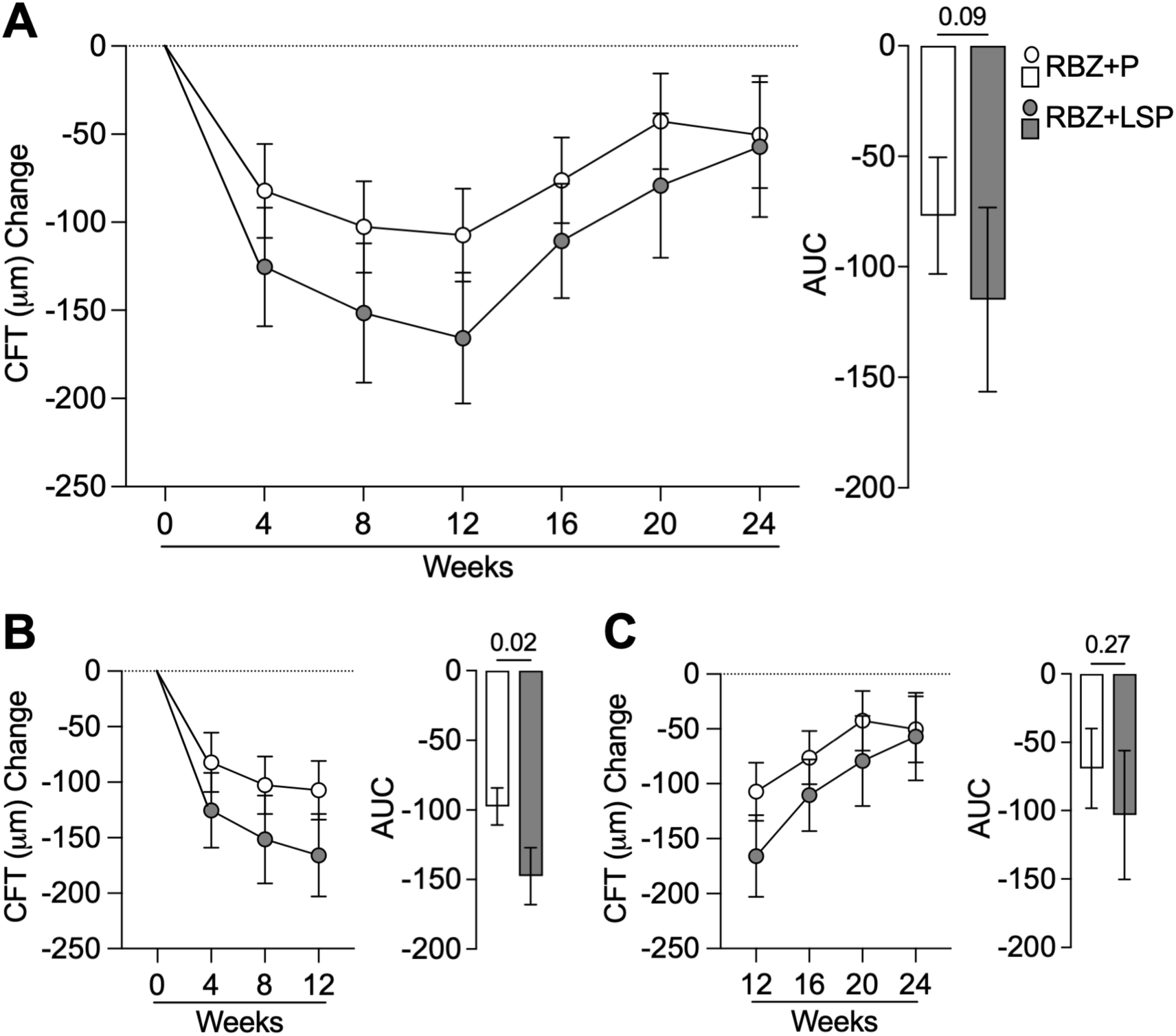
The combination of intravitreal ranibizumab (RBZ) with oral levosulpiride (LSP) improved the change from baseline in central foveal thickness (CFT). Mean longitudinal changes in CFT are shown for the same eyes at baseline (week 0) and after treatment at weeks 4 to 24 **(A)**, weeks 4 to 12 **(B**), and weeks 12 to 24 **(C)**. Data are presented for eyes from patients treated with RBZ combined with either placebo (P) or LSP. Bars represent the area under the curve (AU) comparing the corresponding groups. Values for the first 12 weeks are expressed as mean ± SEM for 18 and 19 eyes in the RBZ+P and the RBZ+LSP groups, respectively; for the last 12 weeks, values are mean ± SEM for 12 and 11 eyes in the RBZ+P and the RBZ+LSP groups, respectively. AUC values were compared using an unpaired t-test, with p values indicated.

Figure 4A illustrates visual and anatomical changes over baseline (week 0) at weeks 12 and 24 in two eyes from different patients in the RBZ+LSP and RBZ+P groups. In each group, an eye is classified as a good responder, showing a gain of more than 5 letters in BCVA and a reduction of more than 50 µm in CFT, and is classified as poor responder, with less than 5 letters gained or 5 or more letters lost in BCVA and a reduction or gain of less than 50 µm in CFT.

**Figure 4.**
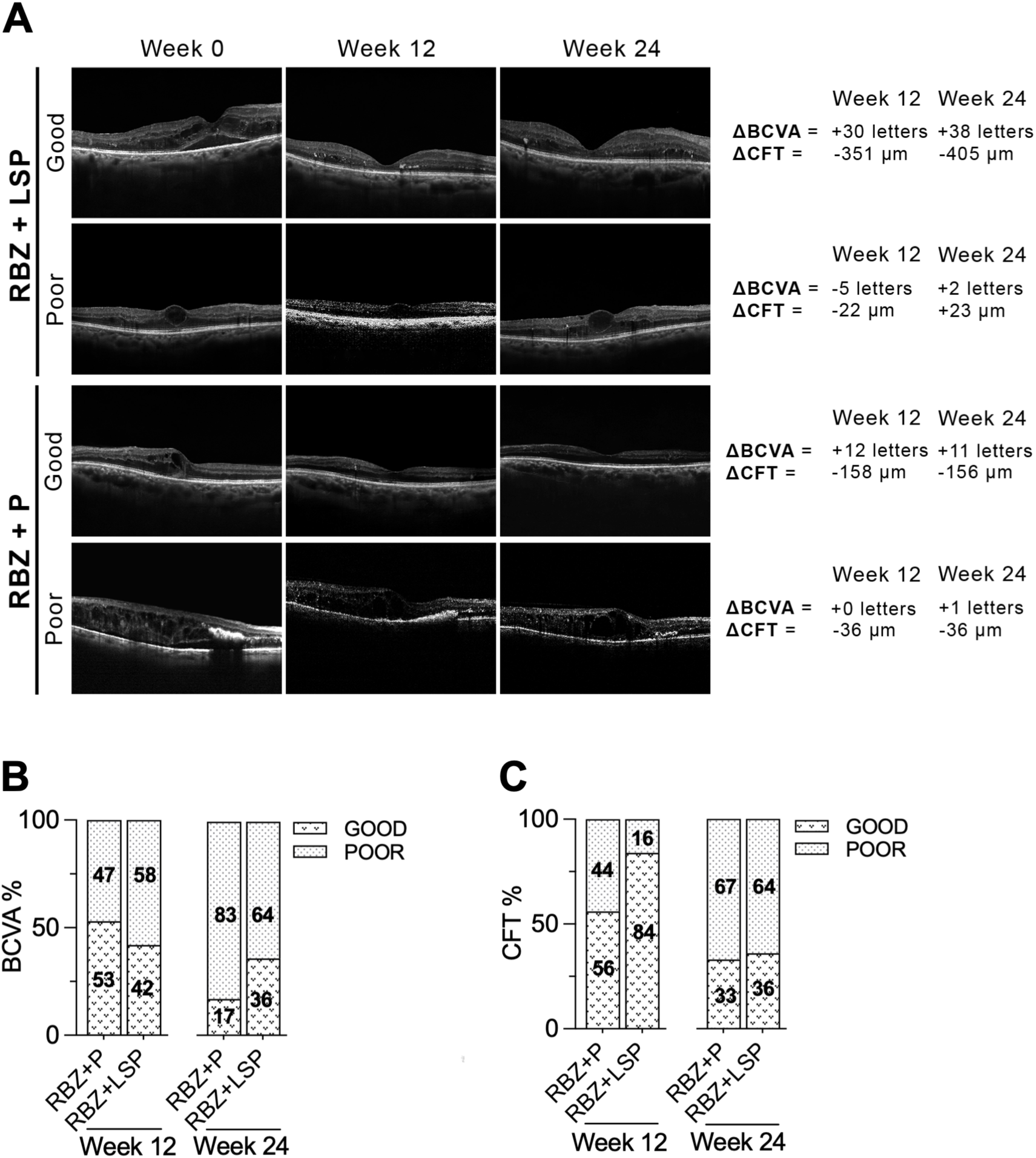
The combination of intravitreal ranibizumab (RBZ) with oral levosulpiride (LSP) increased the proportion of eyes with favourable visual and anatomical responses. **(A)** Representative examples of eyes treated with RBZ combined with either oral LSP or placebo (P) showing changes from baseline (week 0) to weeks 12 and 24 in OCT macular image, best corrected visual acuity (ι1BCVA), and central foveal thickness (ι1CFT). Eyes classified as a good responders demonstrated a gain of more than 5 letters in BCVA and a reduction of more than 50 µm in CFT. Poor responders were defined as eyes with fewer than 5 letters gained or 5 or more letters lost in BCVA and a reduction or increase of less than 50 µm in CFT. The proportion of eyes classified as good or poor responders in BCVA **(B)** and in CFT **(C)** are shown after 12 and 24 weeks of treatment with RBZ combined with either placebo (P) or LSP.

At week 12 the percentage of eyes classified as good responders in BCVA was similar between the RBZ+LSP and the RBZ+P groups (42% vs. 53%) (Fig. 4B), whereas a higher proportion of eyes in the RBZ+LSP group achieved a good response in CFT (84% vs. 56%) (Fig. 4C). By week 24, the RBZ+LSP group showed a greater proportion of good responders in BCVA relative to the RBZ+P group (36% vs. 17%) (Fig. 4B) and the percentage of eyes classified as good responders in CFT was comparable between the RBZ+LSP group and the RBZ+P groups (36% vs. 33%) (Fig. 4C).

Collectively, these results suggest that combining RBZ with LSP produces more rapid early anatomical improvements and sustains greater visual acuity gains over time compared to treatment with RBZ and P.

### Safety

Two patients (11%) in the RBZ-LSP group reported experiencing somnolence, a known side effect of LSP [12]. Somnolence appeared at either 8 or 12 weeks after starting treatment and completely resolved within 4 weeks. There were no significant differences between the RBZ+LSP and the RBZ+P groups in mean serum levels of HbA1c, fasting serum glucose, glomerular filtration rate, or mean intraocular pressure (Table 1).

## DISCUSSION

LSP, a dopamine D2-receptor blocker widely prescribed for diabetic gastroparesis [13], has recently been repurposed as a non-invasive, effective, and safe therapeutic option for naïve patients with centre-involving DMO [4] demonstrating efficacies comparable to RBZ [5]. In this randomized, prospective, placebo-controlled clinical trial, we have investigated the extended therapeutic potential of LSP as an adjunct to intravitreal RBZ in patients with persistent centre-involving DMO.

The combination of LSP with intravitreal RBZ led to faster and longer-lasting benefits compared to the combination of RBZ and P. Both treatment groups showed significant improvements in mean BCVA and CFT values at 12 weeks; however, these improvements were sustained at 24 weeks only in the RBZ+LSP group. Longitudinal analysis of individual eyes before and after treatment using the area under the curve method, revealed superior anatomical and visual outcomes throughout the 24-week period in the RBZ+LSP group. Significant reductions in CFT were noted during the initial 12 weeks, while significant gains in BCVA were maintained during the final 12 weeks of the study. Similarly, the combination of LSP with RBZ increased the proportion of anatomical good responder eyes at 12 weeks and the proportion of eyes with good visual acuity responses at 24 weeks.

The broad pharmacological actions of LSP may underlie its adjunctive anatomical and visual benefits in DMO. Anatomical improvements, indicative of macular oedema resolution, are primarily achieved through the inhibition of specific members of VEGF family. LSP effectively reduces retinal vascular leakage induced by VEGF [7] and downregulates intravitreal levels of both VEGF and PlGF [4], and PlGF is also a key mediator of vascular permeability [14]. Additionally, by blocking dopamine D2 receptors in the anterior pituitary, LSP induces hyperprolactinemia [15], which subsequently elevates ocular vasoinhibin levels −a prolactin-derived fragment known to inhibit vascular leakage caused by VEGF, bradykinin, and diabetic conditions in rodent retinas [6–9,16]. While macular oedema significantly contributes to visual impairment, it is not the sole factor influencing reduced visual acuity, and its resolution does not always correlate with visual improvement [17,18]. In this context, LSP may protect visual function through alternative mechanisms, including those associated with hyperprolactinemia itself. The prolactin receptor is widely expressed throughout the retina [19], and prolactin signalling mitigates photoreceptor cell death and electroretinogram deficits resulting from light-induced retinal damage [20] and aging [21]. Furthermore, LSP’s direct blockage of dopamine D2 receptors in the retina may also enhance vision. Dopamine released by amacrine and interplexiform cells acts as a neuromodulator by influencing retinal circuitry via D1 and D2 receptors across retinal neurons [22], a modulation that can optimize visual processing [23]. Although further research is warranted to fully clarify the mechanisms by which LSP exerts its effects, current evidence supports its potential as a valuable adjunct to first-line intravitreal anti-VEGF therapies in persistent DMO.

Recurrences of macular oedema and the requirement for reinjection remain major challenges in the pharmacological management of DMO. In our study cohort, patients with previously treated recurrent DMO comprised 60% to 77.8% of participants. Among the two pharmacological regimes evaluated (RBZ combined with LSP and RBZ combined with P) 42 to 84% of eyes demonstrated good responses during the initial 12 weeks, while sustained efficacy at 24 weeks was only observed in 17 to 36% of cases. The superior anatomical and visual outcomes associated with the combination of RBZ+LSP, yet the overall need for reinjection remained similar between the two treatments over six months suggesting comparable efficacy in the longer term. It remains to be determined whether the adjunctive use of LSP can reduce the injection frequency in treatment naïve DMO patients compared to those with recurrent DMO following prior therapy.

No significant systemic adverse events were attributed to LSP during the study. Somnolence was reported in two of eighteen patients (11%) at 8 and 12 weeks after treatment initiation, resolving spontaneously within four weeks. Other common side effects associated with LSP, including breast tenderness, reduced libido, or drowsiness, were not observed. This safety profile is consistent with expectations, given that LSP was administered at the prokinetic oral dose of 25 mg three times daily–a dosage previously demonstrated to be safe for long-term treatment of diabetic gastroparesis over periods ranging from 4 to 42 months [12,24,25].

Our study has strengths. It features a prospective, randomized design, which provides robust evidence for treatment efficacy. Adherence to treatment was effectively monitored through the strong indicator of LSP-induced hyperprolactinemia. The inclusion of a placebo group allowed comparison with the natural progression under RBZ monotherapy. Patients and outcome assessors were blinded to the treatment allocation, and the treatment regimens were strictly defined by the study protocol. However, the study has several limitations. The small number of patients in each group may have limited the statistical power of some analyses, although we still observed clinically meaningful and statistically relevant findings. The relatively short follow-up period limits the evaluation of long-term improvements in visual function. Additionally, the heterogeneity of interventions used to treat recurrent DMO prior to study entry could have influenced the outcomes. Lastly, the study protocol had to end prematurely due to shortage of RBZ in Mexico.

In conclusion, our work supports oral LSP as safe effective adjunct to intravitreal anti-VEGF therapies in the management of DMO, particularly in patients with recurrent disease. This approach may provide a more efficient treatment option. However, larger and longer-term studies are required to validate and extend these findings.

## Data Availability

All data produced in the present work are contained in the manuscript.

## ACKNOWLEDGEMENTS

The authors acknowledge the substantial contribution of Nancy Sánchez Martínez and Graciela Ibarra Vargas for the assessment and interpretation of optometry, optical coherence tomography, and fluorescein angiography data; Xarubet Ruíz-Herrera and Fernando López Barrera for data presentation and administration support; and D.A.R. Juan José Díaz Esquivel from Laboratorio de Ensayos de Desarrollo Farmacéutico, Facultad de Estudios Superiores Cuautitlán, UNAM for producing the placebo.

## AUTHOR CONTRIBUTION STATEMENT

EA-C, CDN-A, MLR-O, R G-F, MG-R, EL-S, CC conceived and designed the study; EA-C, CD. N-A, JV, IHI, AH-Q, BER-C, MG-R acquired data; EA-C, IHI, AH-Q, BER-C, MLR-O, GME, CC analysed and interpreted the data; EA-C, CC wrote the manuscript; CC obtained funding; All authors critically revised and approved the final version.

